# How can risk of COVID-19 transmission be minimised in domiciliary care for older people: development, parameterisation and initial results of a simple mathematical model

**DOI:** 10.1101/2021.05.05.21256598

**Authors:** István Z. Kiss, Konstantin B. Blyuss, Yuliya N. Kyrychko, Jo Middleton, Daniel Roland, Lavinia Bertini, Leanne Bogen-Johnston, Wendy Wood, Rebecca Sharp, Julien Forder, Jackie Cassell

## Abstract

This paper proposes and analyses a stochastic model for the spread of an infectious disease that is transmitted between clients and care workers in the UK domiciliary care setting. Interactions between clients and care workers are modelled using specially generated networks, with network parameters reflecting realistic patterns of care needs and visit allocation. These networks are then used to simulate and SEIR-type epidemic dynamics with different numbers of infectious and recovery stages. The results indicate that with the same overall capacity provided by care workers, the minimum peak proportion of infection, and the smallest overall size of infection are achieved for the highest proportion of overlap between visit allocation, i.e. when care workers have the highest chances of being allocated a visit to the same client they have visited before. An intuitive explanation of this is that while providing the required care coverage, maximising overlap in visit allocation reduces the possibility of an infectious care worker inadvertently spreading the infection to other clients. The model is quite generic and can be adapted to any particular directly transmitted infectious disease, such as, more recently, COVID-19, provided accurate estimates of disease parameters can be obtained from real data.

## 1 Introduction

The catastrophic impact of Covid-19 in care homes for older people has been well documented [9, 13], demonstrating the extreme vulnerability of their frail elderly residents to this emerging infection. Less well recognised is the large number of similarly frail older people receiving professional (“regulated”) care in their own homes by visiting careers who visit many clients in different households, known as home care or domiciliary care. In 2020, an estimated 715,000 people provided care to around 330,000 community care users above 65 years old in England [8, 7].

Vacancy in the domiciliary-care-providing sector are typically 1 in 10, with annual staff turnover of 1 in 5 [8]. Social care can contribute to transmission of COVID-19 and other infections such as influenza or norovirus (winter vomiting). Providers may become unable to deliver care if staff are ill or isolating, putting vulnerable people at risk. In care homes, outbreaks are quickly recognised and can have devastating effects, as we have seen during the Covid-19 pandemic [9, 13]. However, outbreaks are harder to detect in domiciliary care, as they takes place in people’s home. There is surprisingly little research on domiciliary care generally [2], and particularly on its role in transmission and prevention of infection.

Transmission of Covid-19 is likely to be higher for people receiving domiciliary care because client households are connected via care workers and there are also connections with carers’ own households. Around one fifth of domiciliary care agencies reported providing care to at least one person with suspected or confirmed Covid-19 [3]. Dispersed across households, and protected from knowledge of each other by the confidentiality required of carers and agencies, domiciliary care associated outbreaks of infection can be hard to detect and are probably under-ascertained. Previous work in domiciliary care has explored how difficult it can be to properly identify and address outbreaks of infectious diseases such as scabies [17] and, more recently, Covid-19 [6].

Given the vulnerability of domiciliary care clients to severe outcomes from Covid-19 and other infections, and the need to protect visiting carers and their families from exposure, there is a need to understand transmission dynamics in this neglected and difficult to study setting. Since the start of Covid-19 pandemic, a large number of mathematical models have looked into disease dynamics from the perspective of evaluating the effectiveness of different non-pharmaceutical interventions [5, 18, 20, 1, 12], and assessing various scenarios of lifting lockdown restrictions [14, 19]. Some work has been done on modelling Covid-19 transmission in the care homes [16], where this disease has been associated with a significant death toll, but so far it has not been studied in a residential care setting. We aim to fill this learning gap by developing and parameterising a simple mathematical model of transmission through domiciliary care, and use it to evaluate the potential impact of different policies for the allocation of carers to clients.

## 2 Mathematical model

We model interactions between clients (people receiving care within their own household) and care workers using a bi-partite network, as shown in Fig. 1. The infection can be passed from care workers to clients, and from clients to care workers. The majority of transmission will be associated with movement of care workers between clients in different households.

**Figure 1:**
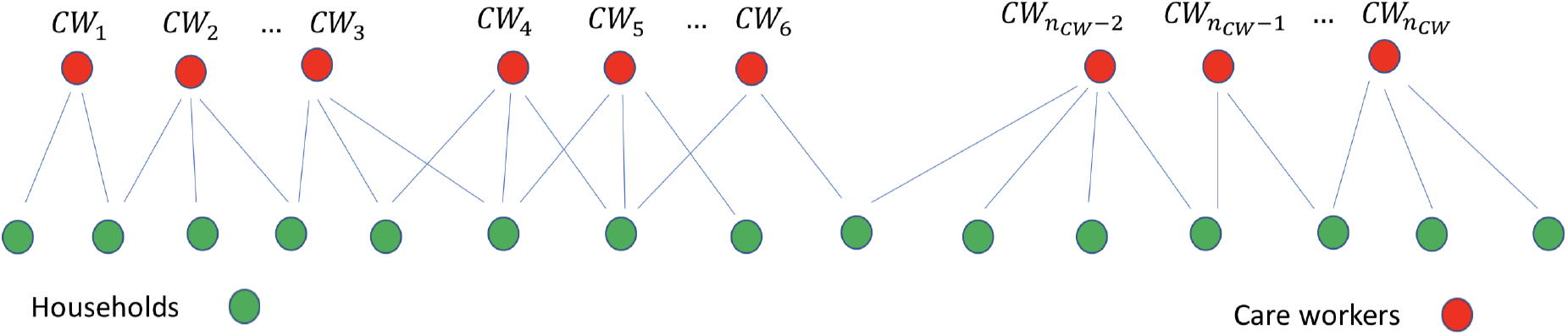
Pictorial representation of the care worker and client/household interactions. Some of these links will be weighted depending on the number of repeat or return visits of care workers to the same client.

The main focus of this study is to understand the impact of care workers’ working pattern on the spread of infectious diseases. In particular we are interested in the difference between whether clients are visited by the same or different care workers. Whilst there is some systematic evidence on the receipt of domiciliary care by clients [11] there is little data on patterns of care delivery. As such we make a number of assumptions based on anecdotal accounts of current practice. The assumptions are as follows:

- Clients require/receive between 1 to 4 visits per day, these can be fulfilled by the same or different care workers
- Care workers are either full-time or part-time with their number of visits per day being around 10 and 5. This approximates the typical 11 to 12 visits per day for full time and 5 to 6 visits per day by part-time care workers. In the former case these are to 7 or 8 different households, while in the latter most visits are to different households.

The networks are generated by allocating stubs to each household. This is done by choosing numbers from a 1 + *Bin*(3, *µ*), with some set value for *µ* such that the mean 1 + 3*µ* is around 3. There are care workers of two types: full-time and part-time. These are also allocate stubs (10 for full-time and 5 for part-time care workers). These stubs are then placed in two separate lists (one for households and one for care workers) with each stub being labelled by the node index (e.g. if node *i* is a household with 3 links the three copies of *i* will be added to the households’ stub list). We then choose stubs at random from the household and from the care worker lists, without replacement. This means that duplicate links are possible and some compatibility conditions need to be observed (i.e. the number of stubs from households has to equal the number of stubs from care workers). Further details are given in the Supplementary Material.

The network has an extra degree of freedom which allows us to vary the amount of repeat visits (later referred to as ‘overlap’). This is done as follows. Once a household and a care worker is connected for the first time, the algorithm searches out all the other stubs from this household and care worker and adds them as extra link with probability *p*_*overlap*_. High values of *p*_*overlap*_ corresponding to the case where care workers go back to the same household, as much as possible. Low values of *p*_*overlap*_ mean visits by care workers are distributed as much as possible and by avoiding repeat visits.

Figure 2 shows the clear difference between the no repeat (left panel) and repeat (right panel) scenarios. We note that thicker edges are weighted from one to four, which reflects the maximum number of visits required by a household. The construction implies that the weight of edges in all three network is the same which means that more overlap between visits leads to a sparser networks, with fewer visible links, but with links of higher weight. This simple models allows us to vary the amount of repeat visits and thus allows us to investigate its impact on infectious disease outbreaks in the domiciliary care sector.

**Figure 2:**
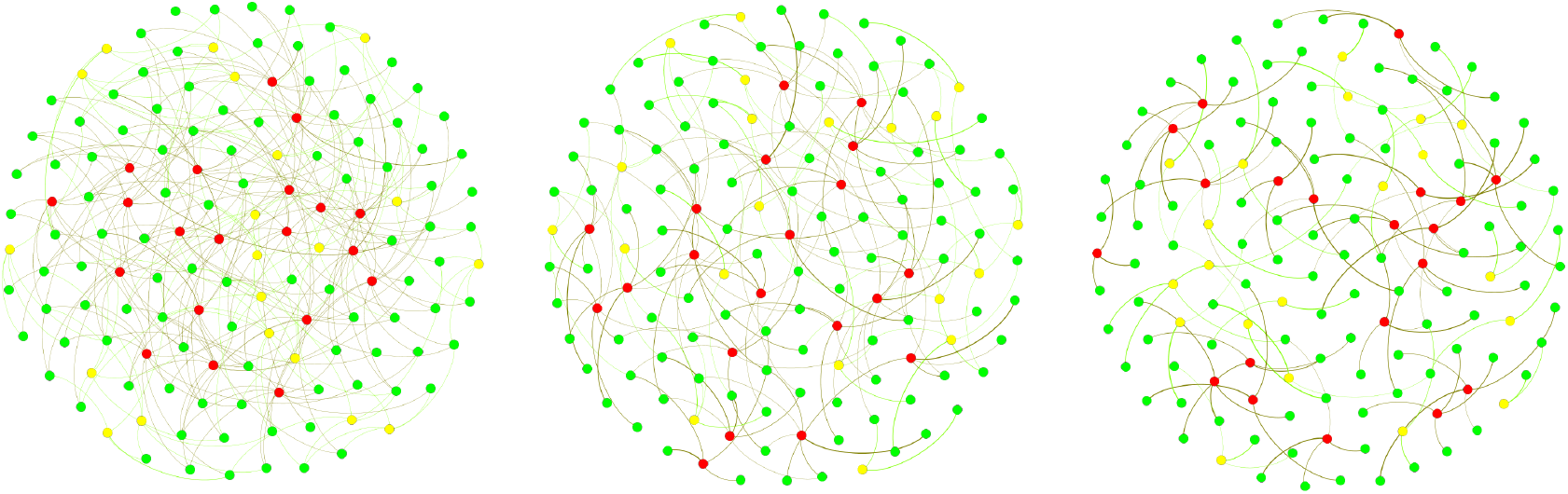
Example of networks with *N*_*HH*_ = 100 households (green nodes), and equal number of full time (red nodes) and part time (yellow nodes) care workers *N*_*FTCW*_ = *N*_*PTCW*_ = 20. *µ* = 2*/*3 in *Bin*(3, *µ*) with *p*_*overlap*_ = 0, 0.5, 1 from left to right.

We are now in a position to impose an epidemic dynamics on the top of the contact structure and we do this by using an *SEIR* model where the nodes can be: susceptible (*S*), exposed (*E*), infected/infectious (*I*) and recovered (*R*). The contact pattern between households and care workers is given by the weighted contact matrix *A* = (*A*_*ij*_)_*i,j*=1,2,…,*N*_, where *N* is the number of nodes in the network. The transitions between different states for each of the nodes are associated with following rates:

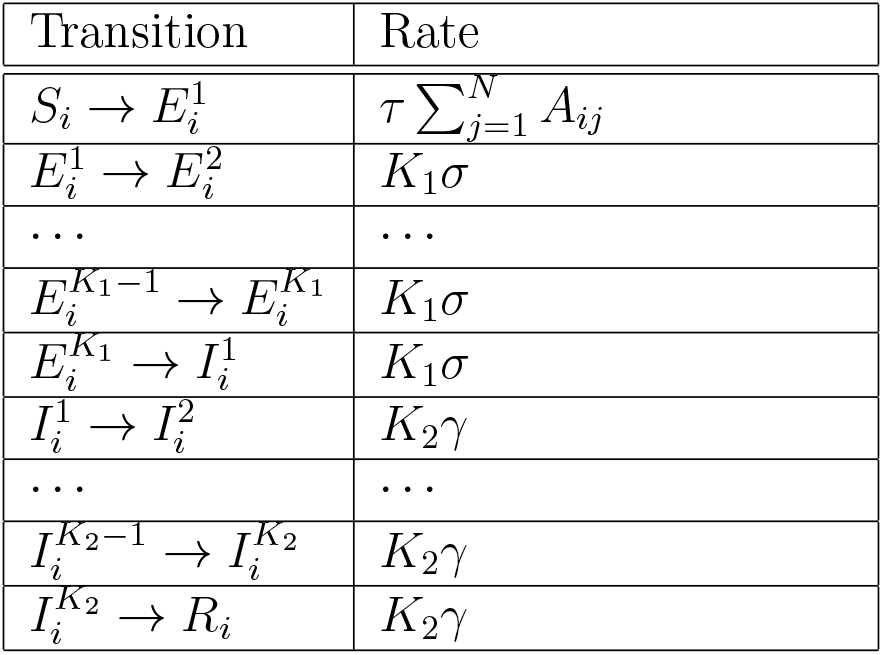

| Transition | Rate |
| --- | --- |
| $S_i \rightarrow E_i^1$ | $\tau \sum_{j=1}^N A_{ij}$ |
| $E_i^1 \rightarrow E_i^2$ | $K_1 \sigma$ |
| … | … |
| $E_i^{K_1-1} \rightarrow E_i^{K_1}$ | $K_1 \sigma$ |
| $E_i^{K_1} \rightarrow I_i^1$ | $K_1 \sigma$ |
| $I_i^1 \rightarrow I_i^2$ | $K_2 \gamma$ |
| … | … |
| $I_i^{K_2-1} \rightarrow I_i^{K_2}$ | $K_2 \gamma$ |
| $I_i^{K_2} \rightarrow R_i$ | $K_2 \gamma$ |

The subscripts *i* = 1, …, *N* are node labels, *τ* is the disease transmission rate per single link in the contact network, 1*/σ* is the incubation period represented by *K*_1_ stages, 1*/γ* is the recovery period represented by *K*_2_ stages. The network is encoded by a weighted adjacency matrix *A* = (*A*_*ij*_)_*i,j*=1,2,…,*N*_.

This model is simulated numerically, using the Gillespie algorithm on the generated contact networks with different overlap.

## 3 Results

To model the dynamics of the infectious spread, we have considered networks going from no overlap to high overlap, see figure 2. In the no overlap (unweighted) network case (see left panel), client needs are met by different care workers, and could potentially involve no return visit by the same care worker. In the high overlap (weighted) network case (see right panel), we assume that if a client is already receiving a visit from a particular care worker and if they require additional visits, then these would most likely be provided by the same care worker.

To analyse the effect of possible overlap on epidemic dynamics, we have performed simulations as follows. Varying the probability *p*_*overlap*_ between 0 and 1 in steps of 0.1, for each value of *p*_*overlap*_, we generated 10 networks, in which weight distribution account for that particular value of *p*_*overlap*_, and then on each of those ten networks, we simulated epidemic dynamics 10 times, thus effectively obtaining 100 simulations for each value of *p*_*overlap*_. These individual simulations were then averaged, and they provided the results shown in Fig. 3, which illustrates how proportions of infected clients, all care workers, and separately full-time and part-time care workers, change depending on *p*_*overlap*_. We observe that increasing the degree of overlap reduces epidemic peaks, which can be explained by the fact the contribution of care workers who make repeat visits to the same household to the overall spread of the infection is limited. Repeat visits to same household make transmission to these households more likely, but this limits the number of links (i.e. opportunities for wider transmission) available for further or wider spread. Fewer links of higher weight thus limit the reach of the epidemic/outbreak, and the higher is the proportion/weight of links connecting the same pairs of care workers and clients, the higher is the reduction in epidemic peak. Figure 4 illustrates this in more detail by showing how the peak in the total proportion of infected individuals, as well as the final epidemic size (overall proportion of initial population who have been infected during the course of epidemic) are both monotonically decreasing with *p*, once again indicating that increasing the degree of overlap between visit allocations reduces the potential for disease spread.

**Figure 3:**
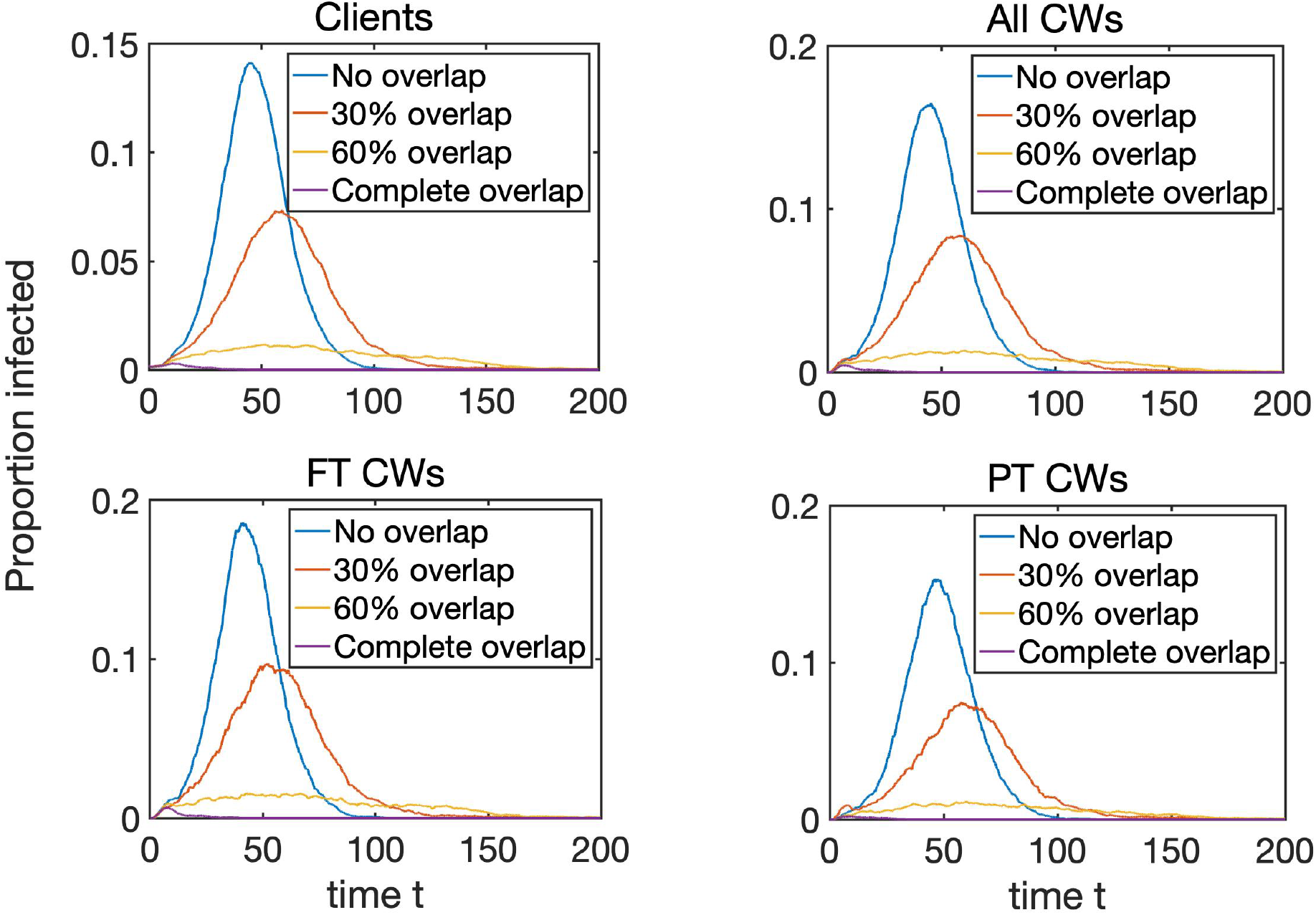
Dynamics of the proportion of infected (exposed *E* and infectious *I*) clients, all care workers, as well as full-time and part-time care workers, depending on the level of overlap *p*_*overlap*_. Epidemics start with one infected node chosen at random, and each trajectory represents an average taken over 10 realisations for 10 networks for each value of *p*. Further parameters are: the rate of infection *τ* = 0.2; the rate of recovery from the *E* state is *s* = 0.3; the rate of recovery from the *I* state is *γ* = 0.3; the number of *E* stages is *K*_1_ = 3; and the number of *I* stages is *K*_2_ = 5. The underlying networks have *N*_*HH*_ = 500 households, and equal number of full-time and part-time care workers, *N*_*FTCW*_ = *N*_*PTCW*_ = 100, with each making *n*_*FTCW*_ *‘*.:: 10 and *n*_*PTCW*_ *‘*.:: 5 visits.

**Figure 4:**
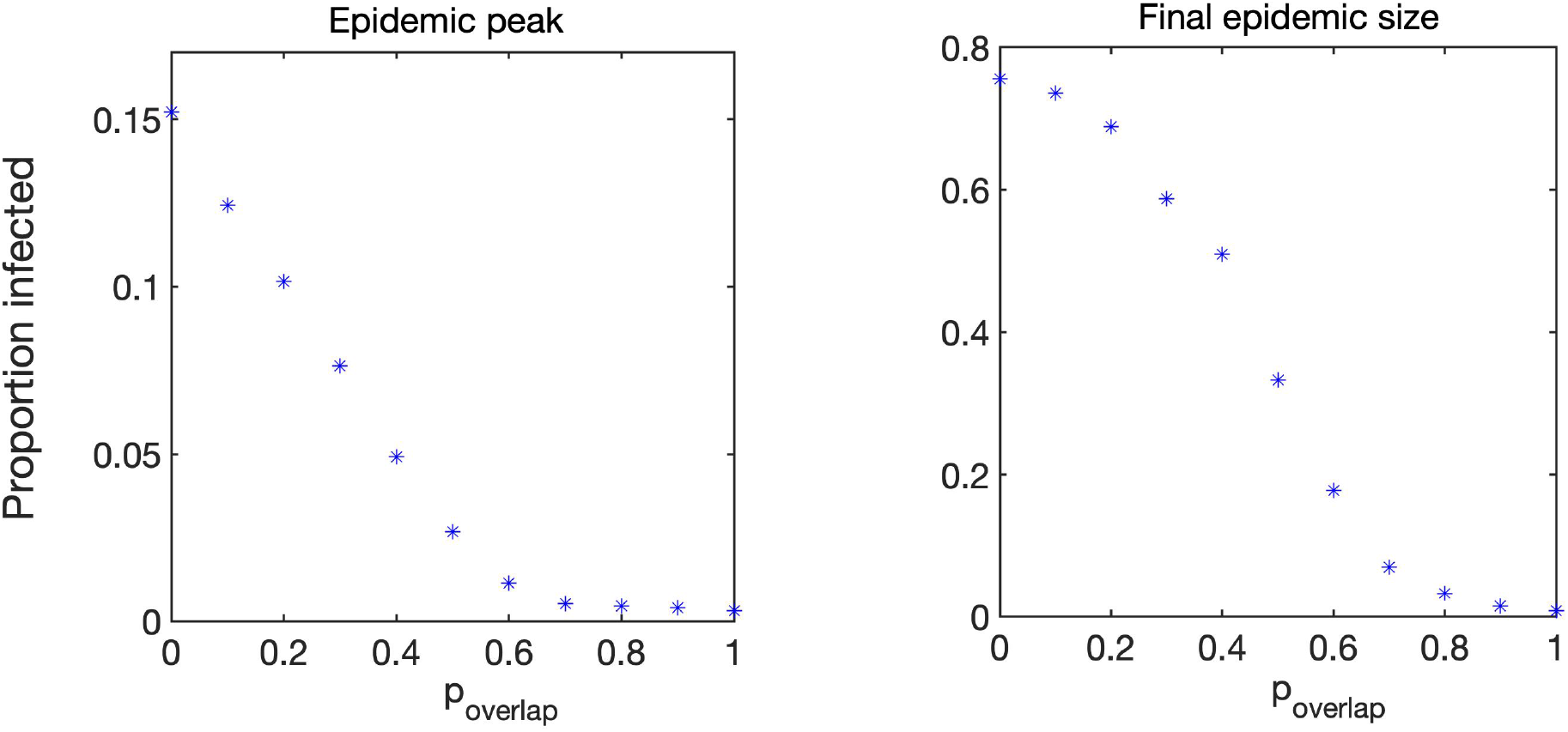
Peak total proportion of infected individuals and final epidemic size.

The networks that we use have the same number of visits, i.e. the sum of weights on all networks are the same, but the number of links is smaller in the weighted networks, since links account for multiple visits. Also, in weighted networks we varied the proportion of overlap between visits of care workers to the same clients to explore the effect this may have on the dynamics.

## 4 Discussion

In this paper, we were able to develop and parameterise a simple model of domiciliary-care-related infectious disease-transmission, with Covid-19 transmission as a test case. The model shows that maximising the number of return visits (i.e. a care worker visiting the same client during the day or multiple days), while fulfilling care needs, has the potential to limit the peak size and the overall burden of a domiciliary care outbreak. This result supports a policy of maximising the ratio of repeat to one-off visits in designing rosters.

There is a strong consensus that care is of higher quality when provided by a small number of familiar and known individuals [10]. However, the extent to which this is possible will be limited by high vacancy rates (8.2%) and the need to ensure adequate remuneration to retain a workforce of which 58% of domiciliary carers are on zero hours contracts and overall 58% of care workers in the independent sector in 2019-20 were paid less than the current national living wage (*£*8.91) [8]. Other limiting factors may include the need of workers to isolate and lose earnings. In the case where such idle time are paid will increase costs, while if unpaid will disincentives workers who will in this case be subsidising the costs of reducing infection risk.

This simple model does not take into account the interaction between the community and household risks of domiciliary care workers. Future models could usefully include this information, which may be important in understanding the force of infection on domiciliary care client households. Lower waged workers typically live in more densely crowded accommodation, and have experienced higher burdens of Covid-19 infection as low socioeconomic conditions and overcrowded housing are risk factor [4, 15]. By contrast domiciliary care clients generally live in small households with few inter-household bridges. We have also not taken into account inter-worker networks, whether household, social or transport related. These too may be important, as domiciliary care workers mostly provide their own transport, which may be shared with families and co-workers. Work transmission has been shown to be an an important contributor to non-household transmission [9].

This simple proof of concept model of domiciliary care provides support from an infection control perspective for the current policy consensus that care should be provided by a limited number of familiar carers. The practical implications of doing so in the context of a setting under considerable staffing and financial pressures need to be carefully considered in the development of any new social care policy. There is a need to extend this model to include the social, household and wider community connections of domiciliary care workers and client households. It should also be extended to address the implications of isolation and contact tracing policies for carers, clients, households and for the sustainability of provision in the face of infection risks. Better data, analysis of patterns of use and delivery of home care services is also a further research need. This will enable policy makers, commissioners and clients to understand the most effective actions and workable trade-offs in relation to risk of Covid-19 and other infections that can be facilitated through domiciliary care, affecting clients, carers and their households. This relatively neglected setting deserves focussed research, enabling clients, carers and their families to be optimally protected.

## Data Availability

All relevant data is listed in the manuscript.

## 5 Supplementary Material

Here we provide some further details for the network construction algorithm.

- Number of households receiving care, *N*_*HH*_;
- Number of full-time (*N*_*FTCW*_) and part-time (*N*_*PTCW*_), giving *N* = *N*_*HH*_ +*N*_*FTCW*_ + *N*_*PTCW*_ nodes in the network;
- Distribution of the number of visits a household or client receives/needs = 1 + *Bin*(3, *µ* = 2*/*3), which then translates to numbers drawn from this distribution, i.e. 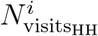, where *i* = 1, 2, …, *N*_*HH*_;
- Number of visits by full-time care workers is *n*_*FTCW*_ and it is *n*_*PTCW*_ for part-time care workers;
- We now allocate a proportion *p*_*FTCW*_ = *n*_*FTCW*_ *N*_*FTCW*_ */*(*n*_*FTCW*_ *N*_*FTCW*_ +*n*_*PTCW*_ *N*_*PTCW*_) of all household stubs to full-time care workers and the remaining ones to part-time care workers. At this point the stubs are not yet allocated to care workers.
- The number of visits made by care workers is now allocated based on 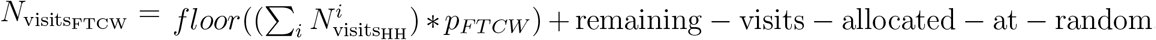, with a similar formula for the part-time care workers. This means that some care workers will have *±*1 or so stubs compared to *n*_*FTCW*_ or *n*_*PTCW*_. But such differences are minimised by a careful choice of the number of care workers of different type;
- Create a list/array by placing copies/labels of *HH* in a list as many times as their number of visits requires;
- Do the same for full-time and part-time *CW* in a different list;
- Pick elements at random from both lists and connect them up;
- This will produce a network where most links have only been realised once, with some duplicate links occurring;
- This is then refined to allow multiple links between the same household and *CW* leading to weighted edges. This is done as follows. Once a *HH* and a *CW* is connected, the algorithm looks for any of stubs belonging to the same *HH* and *CW* and connects them with probability *p*_*overlap*_.

## Acknowledgements

This research, in particular the work of Leanne Bogen-Johnston, Rebecca Sharp, Julien Forder and Jackie Cassell, was supported by the National Institute for Health Research (NIHR) Applied Research Collaboration Kent, Surrey, Sussex. The views expressed are those of the authors and not necessarily those of the NHS, the NIHR or the Department of Health and Social Care.

## Notes

### Competing Interest Statement

The authors have declared no competing interest.

## References

[1] M Aguiar, E M Ortuondo, J B Van-Dierdonck, J Mar, and N Stollenwerk. Modelling covid 19 in the basque country from introduction to control measure response. Scientific Reports, 10:17306, 2020.

[2] National Audit Office: Report by the Comptroller and Auditor General. Adult social care in England: overview. 2014.

[3] Care Quality Commission (CQC). Covid-19 insight. 2020.

[4] Konstantinos Daras, Alexandros Alexiou, Tanith C Rose, Iain Buchan, David Taylor-Robinson, and Benjamin Barr. How does vulnerability to covid-19 vary between communities in england? developing a small area vulnerability index (savi). J Epidemiol Community Health, 2021.

[5] N G Davies, A J Kucharski, R M Eggo, A Gimma, and W J Edmunds. Effects of non-pharmaceutical interventions on covid-19 cases, deaths, and demand for hospital services in the uk: a modelling study. Lancet Public Health, 5:e375–e385, 2020.

[6] Walter D Dawson, Elizabeth C Ashcroft, Klara Lorenz-Dant, and Adelina Comas-Herrera. Mitigating the impact of the covid-19 outbreak: A review of international measures to support community-based care. 2020.

[7] NHS Digital. Adult Social Care Statistics in England: An Overview. 2020.

[8] Skills for Care. The state of the adult social care sector and workforce in England (October 2020). 2020.

[9] Scientific Advisory Group for Emergencies and Public Health England. PHE: Factors contributing to risk of SARS-CoV2 transmission in various settings, 26 November 2020. 2020.

[10] National Institute for Health and Care Excellence. Home care for older people - Quality standard [QS123]. 2016.

[11] Julien E Forder, Juliette Malley, Stacey Rand, Florin Vadean, Karen C Jones, and Ann Netten. Identifying the impact of adult social care: interpreting outcomes data for use in the adult social care outcomes framework. 2016.

[12] G Giordano, F Blanchini, R Bruno, P Colaneri, A Di Filippo, A Di Matteo, and M Colaneri. Modelling the covid-19 epidemic and implementation of population-wide interventions in italy. Nat. Med., 26:855–860, 2020.

[13] Ian Hall, Lorenzo Pellis, Thomas House, Hugo Lewkowicz, James Sedgwick, and Nick Gent. Rapid increase of care homes reporting outbreaks a sign of eventual substantial disease burden. medRxiv, 2020.

[14] M J Keeling, G Guyver-Fletcher, A Holmes, L Dyson, M J Tildesley, and E M Hill. Precautionary breaks: Planned, limited duration circuit breaks to control the prevalence of covid-19. SSRN, medRxiv, 2020.

[15] Nazir I Lone, Joanne McPeake, Neil I Stewart, Michael C Blayney, Robert Chan Seem, Lorraine Donaldson, Elaine Glass, Catriona Haddow, Ros Hall, Caroline Martin, et al. Influence of socioeconomic deprivation on interventions and outcomes for patients admitted with covid-19 to critical care units in scotland: A national cohort study. The Lancet Regional Health-Europe, 1:100005, 2021.

[16] L Nguyen, S Howick, D McLafferty, G Anderson, S Pravinkumar, R Van Der Meer, and I Megiddo. Evaluating intervention strategies in controlling coronavirus disease 2019 (covid-19) spread in care homes: An agent-based model. Infection Control & Hospital Epidemiology, pages 1–11, 2020.

[17] Emily Phipps, Maaike E Pietzsch, Jackie A Cassell, and Clare Humphreys. The public health importance of scabies in community domiciliary care settings: an exploratory cross-sectional survey of health protection teams in england. Epidemiology & Infection, 147, 2019.

[18] H Sjödin, A F Johansson, and et al. Brännström, A. Covid-19 healthcare demand and mortality in sweden in response to non-pharmaceutical mitigation and suppression scenarios. International Journal of Epidemiology, pages 1443–1453, 2020.

[19] R N Thompson, T D Hollingsworth, V Isham, and et al. Key questions for modelling covid-19 exit strategies. Proc. Roy. Soc. B, 287:20201405, 2020.

[20] A R Tuite, D N Fisman, and A L Greer. Mathematical modelling of covid-19 transmission and mitigation strategies in the population of ontario, canada. CMAJ, 192:E497–E505, 2020.

